# THE THERAPEUTIC POTENTIAL OF IVERMECTIN FOR COVID-19: A SYSTEMATIC REVIEW OF MECHANISMS AND EVIDENCE

**DOI:** 10.1101/2020.11.30.20236570

**Authors:** Stefanie Kalfas, Kumar Visvanathan, Kim Chan, John Drago

## Abstract

**Introduction:** Ivermectin is a commonly used antihelminthic agent with over 35 years of established safety data in humans. Recent data demonstrates antiviral activity in vitro against SARS-CoV-2, in addition to a range of viruses. In vitro and animal models also provide evidence of immunomodulatory action. These additional modes of action are supported by in silico modelling, which propose a number of viral and host targets that would mediate these effects.

**Objectives:** The aim of this study is to systematically review the published and preprint clinical literature and study results that assessed the potential role of ivermectin as a COVID-19 therapeutic and prophylactic agent.

**Methods:** We conducted a comprehensive review of PubMed, medRxiv, ClinicalTrials.gov, Global Coronavirus COVID-19 Clinical Trial Tracker, World Health Organization International Clinical Trials Registry Platform, EU Clinical Trials Register, ANZ clinical trials registry, and references from relevant articles.

**Results:** Search keywords- “COVID-19 (and synonyms) AND ivermectin”- generated 86 articles on PubMed, 48 on medRvix and 37 on clinicaltrials.gov at the time of writing. Twelve of these were listed as completed clinical trials and of these, 8 were included as investigators had released results. Positive mortality benefit, reduced time to clinical recovery, reduced incidence of disease progression and decreased duration of hospital admission were reported in patients across all stages of clinical severity.

**Limitations:** Due to the time-critical nature of the COVID-19 pandemic our review included preprint data, which must be interpreted with caution while it awaits peer review.

## INTRODUCTION- COVID-19

The health and economic impact of COVID-19 is unparalleled. There has been no threat in recent times of the magnitude of COVID-19 to human survival and economic stability, with over 1,190,000 deaths reported globally to date and baseline economic forecasts of a 5·2 % contraction in global GDP in 2020.^1,2^ The likelihood of death in an at-risk population is substantial, with an estimated infection fatality ratio ranging between 11·6% and 16·4% in men ≥80 years, and between 4·6% and 6·5% in women ≥80 years.^3^ There is no validated vaccine for COVID-19 and there is currently no approved drug therapy that when given early in the disease course reduces morbidity or mortality. Treatments must therefore be established to mitigate the current global crisis. Furthermore, in order for these to be delivered in an egalitarian fashion, these must be affordable with scalable manufacturing potential to allow for delivery of therapy to be equitable and widespread. Repurposing of the anti-parasitic drug ivermectin to treat COVID-19 has become a point of scientific investigation following a study in Australia, which demonstrated its efficacy against SARS-CoV-2 in vitro.^4^ As many countries around the world find themselves in the grips of progressively more ominous second and third epidemic waves, we present the current in vitro, in silico and in vivo data surrounding the potential role of ivermectin in treating COVID-19.

## IVERMECTIN- AN OVERVIEW

Ivermectin is a broad spectrum anti-parasitic agent that was first semi-synthetically derived from a fermentation broth of the soil bacteria Streptomyces avermitilis in 1975 in Japan.^5,6^ It belongs to the avermectin family and is the most widely used anti-parasitic in this class of drugs.^6^ It is listed as an essential medication by WHO and has been called a “wonder drug”.^7^ In 2015, William Campbell and Satoshi Ōmura were awarded a joint Nobel Prize in Medicine for their discovery and development of ivermectin.

For over 35 years ivermectin has been successfully used to treat various parasitic infections in humans and animals. Hundreds of millions of courses of ivermectin are delivered every year through mass drug administration campaigns as well as on an individual basis for the eradication of helminthic and arthropodal infections. Ivermectin binds to glutamate gated chloride channels in invertebrate nerve and muscle cell membranes, resulting in membrane hyperpolarisation which then leads to paralysis and death. These chloride channels are specific to protostome invertebrate phyla.^8^ While they are closely related to mammalian glycine receptors, ivermectin has a low affinity for mammalian ligand-gated chloride channels. In addition, ivermectin does not readily cross an intact blood brain barrier.^7^ Mutations in the ABCB1 transporter gene known to occur in some animal species have been reported in humans but are predicted to be rare and should be readily recognized clinically as acute central nervous system (CNS) toxicity.^9^ These signs and symptoms can include tremor, myoclonus, ataxia, drooling, bradypnoea, anorexia, somnolence, mydriasis, salivation and paralysis.

Ivermectin is currently licensed for oral and topical use in humans, and oral, topical and parenteral use in animals, with standard dosing used typically between 150-400 μg/kg.^7^ There a several case reports of ivermectin being successfully used subcutaneously to treat patients with disseminated strongyloidiasis who fail oral therapy.^10^

Ivermectin has a well characterized wide safety margin, with several phase 1 studies demonstrating its safety even at 5-10 times its usual dose of 150-200 μg/kg, although there is limited data in pregnancy.^7,11-15^ There are no absolute drug contraindications listed by the manufacturer.16 There have been rare reports of increased INR when ivermectin has been co-prescribed with warfarin.^17^.

Side effects are usually mild and self-limited, and include headache, dizziness, skin irritation and nausea.^17^ A cumulative incidence of one serious adverse event per million was found over the first 11 years of mass global ivermectin (trade name, Mectizan) administration ^18^

## IVERMECTIN- ANTIVIRAL PROPERTIES

In addition to its anti-helminthic effects, ivermectin has also been shown to have antiviral activity in vitro against numerous RNA and DNA viruses, including simian virus 40, pseudorabies virus, human Immune-deficiency virus, dengue virus, West Nile virus, Venezuelan equine encephalitis, influenza virus and yellow fever virus.^19-25^ This broad-spectrum antiviral activity is thought to be a result of the fact that ivermectin inhibits viral protein transport mediated through the host importin α/β heterodimer (IMP-α/β).^24^ Viral protein translocation into the host nucleus through IMP α/β is known to be a crucial aspect of robust infection for many viruses.^26-28^

The virological efficacy of ivermectin against dengue infection has been demonstrated in a phase III clinical trial using 400 μg/kg, although no clinical efficacy was demonstrated in this study.^29^ This may be due to timing and dosing regimen of ivermectin.

An in vitro study recently demonstrated that ivermectin is active against SARS-CoV2, with concentrations of 5 μM resulting in a 99·8% reduction of cell-associated SARS-CoV-2 RNA in 48 hours.^4^ While these drug concentrations are unlikely to be achieved in humans through currently approved oral dosing regimens, this does not discount the possibility of efficacy in vivo for many reasons. First, viral loads typically deployed for in vitro transfection experiments are very high and may not be representative of the clinical scenario. Second, a simple monolayer of the African green monkey kidney cells (Vero-hSLAM cells) with virus introduced in the supernatant does not replicate the complex/dynamic human tissue structure. Third, Vero-hSLAM cells may not be representative of the human model of infection as these cells are not respiratory cells and do not have ACE-2 receptors through which SARS-CoV-2 mediates cell entry.^30^ Fourth, viral replication efficiency may vary in different cell types. Indeed, Vero-hSLAM cells is a cell line favoured specifically for viral replication. Fifth, synergistic effects inherent in the immunomodulatory and anti-inflammatory properties of ivermectin may also result in lower concentrations required to treat COVID-19. Finally, concentration limitations may potentially be overcome through novel routes of administration and dosing regimens.

## IVERMECTIN- ANTI-INFLAMMATORY AND IMMUNOMODULATORY PROPERTIES

Ivermectin and avermectin have been demonstrated to have anti-inflammatory and immunomodulatory actions in several in vitro and animal models, and is licensed for topical use in humans for the treatment of inflammatory lesions in rosacea.^31^

A recent proteomic analysis showed that ivermectin decreases levels of proteins associated with SARS-CoV-2 in an in vitro culture system.^32^ An in vitro model of lipopolysaccharide (LPS) induced inflammation demonstrated that avermectin, from which ivermectin is derived, significantly impairs pro-inflammatory cytokine secretion (interleukin-1 β and tumour necrosis factor-α) by 30% and doubles secretion of the immunoregulatory cytokine interleukin (IL)-10. This effect is thought to be mediated through inhibition of nuclear translocation of nuclear transcription factor κ-B (NF-κB), and phosphorylation of mitogen activated protein (MAP) kinases.^33^ Mice treated with ivermectin demonstrated improved survival rates with a reduction in tumour necrosis factor-α and IL-1, IL-6 compared to controls following a lethal dose of LPS.^34^ The dose used in this study was approximately equivalent to about 18 mg in humans.^35^ In a mouse model of allergic asthma, bronchoalveolar lavage fluid in oral ivermectin treated mice (2 mg/kg) demonstrated a significantly reduced number of immune cells and production of pro-inflammatory cytokines and antibodies compared to controls.^36^ A suppression of mucous secretion by goblet cells was also observed. Immunomodulatory and anti-inflammatory effects have also been observed in a murine model of atopic dermatitis, where ivermectin was shown to improve allergic skin inflammation.^37^ Ivermectin was shown to directly inhibit antigen-specific and non-specific-CD4+ and CD8+ T cell proliferation and effector functions (IL-4, interferon-γ and granzyme B production), whilst having no effect on dendritic cell migration and maturation.

## IVERMECTIN- IN SILICO MODELLING

In silico models have raised the possibility that ivermectin may have several mechanisms of action against COVID-19 in addition to the established inhibition of IMP α/β. It is possible that ivermectin may be able to prevent cell entry of SARS-CoV-2 through blockade of a high affinity docking site on the human angiotensin converting enzyme 2 (ACE-2) receptor that has been identified through two independent computational modelling studies.^38,39^ The positioning of this identified binding site could theoretically interfere with the SARS-CoV-2 spike glycoprotein binding, and thus reduce viral entry into cells. Using a novel computation method to analyse kinetically active residues, Perisic showed homology between the binding site of ivermectin with its known target in parasites (glycine receptor α 3) and proteins on the surface of SARS-CoV-2.^40^ Furthermore, two computational biology and molecular docking studies identified ivermectin as a potentially effective inhibitor of the RNA-dependent RNA polymerase of SARS-CoV-2.^41,42^

## METHODS

We conducted a comprehensive review of PubMed, medRxiv, ClinicalTrials.gov, Global Coronavirus COVID-19 Clinical Trial Tracker, World Health Organization International Clinical Trials Registry Platform, EU Clinical Trials Register, ANZ clinical trials registry, and references from relevant articles. Search keywords were “COVID-19 (and synonyms) AND ivermectin”. SK conducted the systematic review and selected articles. Decision regarding inclusion was then verified by JD and KC. Studies were included if they had released results at the time of writing. Studies were excluded if they had not released results, or if they had been retracted. Studies were not excluded based on site of study, time of follow-up or comparator arm. Study participants were limited to PCR confirmed COVID-19 patients and close contacts (healthcare workers and household contacts) of PCR confirmed COVID-19 patients. The intervention was ivermectin as either a monotherapy or combination therapy for COVID-19 or prophylaxis for COVID-19, irrespective of dose and timing of administration. Outcome measures in the treatment trials included all-cause mortality and time to clinical recovery, in addition secondary measures that included time to viral clearance and disease progression. The primary outcome measure in the prophylaxis trials was development of symptoms consistent with COVID-19 and/or development of PCR confirmed COVID-19.

## RESULTS

Our trials database search using the keywords “COVID-19” and synonyms AND “ivermectin” revealed 86 articles on PubMed, 48 on medRvix and 37 on clinicaltrials.gov at the time of writing. Of these, 12 were completed clinical studies that examined the effect of ivermectin as a combination or single therapy for COVID-19 treatment and prophylaxis. Eight of these had released results at the time of writing this review and met inclusion criteria. Of these studies, four are randomised controlled trials, one is a prospective cohort study, one is a matched case control study, one is a retrospective analysis, and one is a pilot study. Five of these studies were disease treatment trials, and three were prophylaxis trials. We expand upon these published and preprint results below, which are summarised in Table 1 and Table 2.

**Table 1.** Summary of key therapeutic ivermectin trials. (IVM ivermectin. DOXY doxycycline). Ivermectin was administered orally in all trials. “Standard therapy” differed between trials and is defined as the standard therapy administered in the region/hospital at the given period of the trial.

| Study reference | Study design | Study intervention | Total number of participants | Mortality rate (treatment group vs. comparator) | Disease progression or viral clearance (treatment group vs. comparator) | Duration of hospital admission or time to recovery (treatment group vs. comparator) |
| --- | --- | --- | --- | --- | --- | --- |
| Rajter et al. 2020 <sup>42</sup> | Multicentre retrospective analysis | 1-2 doses of 200 µg/kg IVM + standard therapy vs. standard therapy | 280 hospitalised patients<br>(IVM)= 173<br>(control)= 107 | Overall study results<br>15% vs. 25·2% (p=0·03)<br>Severe disease<br>38·8% vs. 80·7%<br>(p=0·001) | No difference in successful rate of extubation | No significant difference |
| Gorial et al. 2020 <sup>45</sup> | Matched case-control | 1 dose of 200 µg/kg IVM + standard therapy vs. standard therapy | 87 hospitalised patients<br>(IVM) = 16<br>(control)= 71 | 0% vs. 2·8%<br>(p value not given) | Time to viral clearance: 7 days (95% CI 6-11) vs. 12 days (95% CI 10-15 ) (p<0·001) | 7·62 ±2·75 vs. 13·22 ±0·90 days (p=0·00005) |
| Carvallo et al. 2020 <sup>46</sup> | Prospective single cohort study (simultaneous untreated controls) | IDEA protocol:<br>Mild cases: IVM 300 µg/kg + aspirin<br>Moderate cases: IVM 450 µg/kg + aspirin + dexamethasone<br>Severe cases: 600 µg/kg IVM + enoxaparin + dexamethasone | 167 mild-severe patients<br>(mild) = 135<br>(moderate) = 12<br>(severe) = 22 | Overall: 0·59%<br>Mild cases: 0%<br>Mod-severe cases: 3·1%<br>Mortality of patients not on IDEA protocol at same hospital over duration of study: 25% | Disease progression:<br>Mild cases: 0% 7 day symptom progression<br>0% required hospitalisation<br>Moderate-severe cases: 1 patient (3·1%) had disease progression | Not reported |
| NCT04523831<sup>50</sup> | Double blind randomised controlled trial | 6-12 mg IVM + 5 days DOXY + standard therapy vs. standard therapy | 363 mild - moderate (treatment) = 183 (control) = 180 | 0 % vs. 1.6% (p value not reported) | Clinical deterioration: 8.7% vs 17.8% (p=0.013)<br>Viral clearance within 12 days: 92.3% vs. 80 % (p value not reported) | Clinical recovery within 7 days:<br>60.7% vs. 44.4% (p=0.03) |
| Hashim et al. 2020<sup>49</sup> | Randomised controlled trial | 2-3 days of 200 µg/kg IVM + 5-10 days DOXY + standard therapy vs. standard therapy | 140 mild-critical patients (treatment) = 70 (48= mild-mod, 11= severe, 11= critical) (control) = 70 (48= mild-mod, 22=severe) | Mild=mod: 0% vs 0%<br>Severe: 0% vs. 27.27 % (p=0.052) | Disease progression in severe cases: 9% vs. 31.81 % (p=0.15) | Time to recovery:<br>10.61±5.3 days vs. 17.9±6.8 days (p<0.01) |

**Table 2.** Summary of key COVID-19 prophylaxis ivermectin trials. (IVM ivermectin. PEE personal protective equipment)

| Study reference | Study design | Study intervention | Total number of participants | Incidence of COVID-19 infection (treatment vs. control) |
| --- | --- | --- | --- | --- |
| NCT04425850<sup>50</sup> | Prospective placebo controlled trial | 1 drop IVM buccal drops (6mg/ml) + 5 sprays carrageenan nasal spray (0.17mg/spray) both repeated 5 times per day + PPE vs. PPE only | 229 Healthcare workers (treatment) = 131 (control) = 98 | 0% vs. 11.22%<br>Positive SARS-CoV2 PCR within 28 days (p<0.0001) |
| NCT04422561<sup>50</sup> | Randomised controlled trial | IVM 200-400 µg/kg oral on day 1 and day 3 vs. no intervention | 304 household contacts of confirmed COVID-19 case | 7.4% vs. 58.4% symptomatic within 14 days (p value not reported) |
| Behera et al. 2020<sup>56</sup> | Matched case-control study | Two doses IVM 300 µg/kg oral + PPE vs. PPE only | 186 matched case-control pairs of Healthcare workers<br>77 controls took IVM<br>38 cases took IVM | IVM use associated with a 73% reduction.<br>OR, 0.27<br>(95% CI 0.15-0.51) |

## DISCUSSION- DATA FROM CLINICAL STUDIES

A multicentre retrospective analysis, known as the ICON study, reported an overall mortality benefit (p=0·03) in moderate-severe COVID-19 patients (n=173) who were given at least one dose of oral ivermectin in addition to standard therapy, compared to matched controls (n=107).^43^ Two hundred and eight consecutive COVID-19 patients recruited across four hospitals in Florida were included in the study undertaken between March 15 and May 11, and divided into a treatment arm and a control arm. Standard approved dosing of 200 μg/kg was used, administered either as a single dose or followed by a second dose on day 7 at the discretion of the physician. Thirteen of the 173 patients who received ivermectin were given a second dose. Most patients in both groups were also prescribed hydroxychloroquine, azithromycin or both, with a higher use of both drugs noted in the control group before the secondary matched analysis. A statistically significant association between reduced mortality and ivermectin use was observed (p= 0·03). This was even more pronounced in the subset of severe patients, with a mortality of 80·7% in controls vs 38·8 in the ivermectin group (p= 0·001). This association remained significant following multivariate analysis to adjust for comorbidities and differences between groups, and in a propensity score-matched cohort (p=0·045 for total group; p= 0·002 in severe subset). The absolute risk reduction was 11·7%. These results compare with the observed absolute risk reduction of dexamethasone use in severe patients of 12·1% seen in the RECOVERY trial, which was pivotal in introducing corticosteroids as standard of care in severe and critical COVID-19 patients in many countries.^44,45^ Despite a significant difference in overall mortality, and mortality in severely unwell COVID-19 patients, the ICON study was not powered to detect a mortality difference in moderately unwell patients, nor a reduction in duration of hospital stay or the rate of successful extubation. As a retrospective analysis it is also limited by potential unmeasured confounding factors, however the ivermectin group tended to have a greater proportion of patients with known COVID-19 risk factors.

A reduced time to hospital discharge was demonstrated in a pilot prospective preprint study of 16 hospitalised COVID-19 patients administered one dose of 200 μg/kg ivermectin in addition to standard therapy, compared to 71 matched controls receiving standard therapy alone (7·62 ±2·75 versus 13·22 ±0·90 days, p=0·00005).^46^ Standard therapy was defined as hydroxychloroquine and azithromycin in this study. A signal for reduced mortality was also observed in the ivermectin group, however the study was not powered to address this endpoint.

A preprint observational analysis in medRxiv has reported on the mortality benefit of ivermectin combination therapy, known as the IDEA protocol, in COVID-19 patients (n=167) when compared to overall COVID-19 mortality in the region across the same time period (p=0·0475).^47^ The IDEA protocol consists of three permutations of an ivermectin combination treatment regime based on severity of COVID-19 upon commencement. This includes incrementally increasing doses of ivermectin and either aspirin or enoxaparin depending on severity of illness, as well as dexamethasone for moderate-severe hospitalised patients. Ivermectin doses of 300 μg/kg, 450 μg/ kg and 600 μg/ kg were prescribed to mild (n=135), moderate (n=12) and severe (n=22) cases respectively on days 0 and 7. A total of 167 patients were included in this study, with an average age of 55·7 years. Primary outcomes were progression of disease and 30 day mortality. There was a 0% rate of symptom progression within 7 days of follow-up in the mild patient group (n=135). No patients in this subgroup required admission to hospital and all made a full recovery. In the moderate to severe group (n=32), only one patient died after both 30 days and 2 months of follow up. Aside from this patient, disease progression did not occur in any of the remaining 31 moderate-severe patients after commencement of the IDEA protocol, and all proceeded to recover. This is in contrast to the interim report from the World Health Organisation (WHO) SOLIDARITY trial, which failed to show a reduction in disease progression with remdesivir, lopinavir, interferon β1 or hydroxychloroquine.^48^ The overall mortality reported with the IDEA protocol was 0·59%. This compares to an overall mortality in the region of approximately 2·1% (p=0·045). The mortality rate in moderate-severe patients (n=32) receiving the IDEA protocol was 3·1%, as compared to a mortality rate of 25% in patients admitted to the same hospital over the same period who did not receive the IDEA protocol (n=12). This study is limited by the absence of a concurrent control arm with a matched number of moderate to severe patients. The overall 28 day mortality in a large randomised placebo controlled trial of remdesivir for COVID-19 was reported as 11·4% in the remdesivir group, compared to 15·2% in the control group.^49^ In the subset of moderate to severe patients (ordinal score 5-7 at baseline) randomised to receive remdesivir, 12·2% died by day 28. The classification of moderate to severe disease between the IDEA and Remdesivir trials was comparable according to the study methods.

Doxycycline is a broad-spectrum antimicrobial with anti-inflammatory activities that has recently been shown to inhibit SARS-CoV-2 cell entry and replication in vitro, leading to trials that examine the synergistic effects of doxycycline and ivermectin.^50,51^ There are now two randomised controlled trials comparing ivermectin and doxycycline combination therapy with standard therapy that have published results on medRxiv and ClinicalTrials.gov (NCT04523831). ^52,53^ The largest of these included 363 mild to moderate COVID-19 patients and tested a treatment regimen of a single dose of 6-12 mg ivermectin on day 0, with a 5 day course of 100mg doxycycline twice daily (NCT04523831).^53^ 183 of the participants were randomised to receive ivermectin combination therapy plus standard care, while the remaining 180 were randomised to receive standard care. Results published on ClinicalTrials.gov indicate a higher rate of early clinical improvement within 7 days in the treatment group (60·7%) compared to control group (44·4%) (p=0·03) in addition to a lower rate of clinical deterioration in the treatment group (8·7%) vs control (17·8%) (p=0·013). This was mirrored in improved rates of viral clearance in the treatment group, where 7·7% of the treatment group vs 20% of controls were still returning a positive SARS-CoV-2 PCR 12 day after initial diagnosis. A mortality signal was also demonstrated, with zero deaths in the treatment group, compared to 3 deaths in the control group (1·6%). This is in contrast to corticosteroids, which, when used early are associated with possible harm.^44,45^ We await final analysis of these results in both preprint and peer-reviewed publication.

A randomised control trial in medRxiv studied the effect of ivermectin and doxycycline combination therapy on severe and critical COVID-19 patients in addition to mild to moderate cases.^52^ Seventy patients were randomised to the treatment arm, which consisted of 2-3 days of 200 μg/kg of ivermectin and 100 mg doxycycline twice daily for 5-10 days, in addition to standard therapy. Forty eight patients in the treatment group were classified as mild-moderate, 11 as severe and 11 critical. This compared to 48 mild-moderate patients in the control arm and 22 severe patients. No critically ill patients were randomised to the control arm due to ethical considerations. A mortality benefit was demonstrated in the severe subset of patients, with a mortality rate of 0% in the treatment group compared to 27·27% in the control group (p=0·052). No patients died in the mild-moderate subsets of either group. The mortality rate in the critical group was 18·2%, and although there was no direct comparator group in this trial, this was lower than the mortality rate of the severe control group. Published mortality rates in COVID-19 patients admitted to ICU range between approximately 25·7%-59·5%.^54^ The WHO SOLIDARITY trial, in which 11,266 adult patients with COVID-19 were randomised to one of 4 or local standard of care reported an mortality rate of 49% amongst critically patients who were already intubated at time of randomisation.^48^ A reduced median time to recovery was also demonstrated in both mild-moderate (6·34±2·4 days vs 13·66±6·4 days) and severe groups (20·27±7·8 vs 24·25±9·5 days) compared to controls, although this only reached statistical significance in the mild-moderate group (p<0·01), possibly due to small sample size of the severe group. This translates to a reduced time to recovery of 7·32 days in the mild-moderate treatment group and 3·98 days in the severe treatment group. The overall median time to recovery in the treatment group was 10·61±5·3 days vs 17·9±6·8 days (p<0·01). Remdesivir has been reported to reduce median time to recovery by 5 days (p<0·001).^55^ This has been conflicted by interim reports from the SOLIDARITY trial, which did not find a significant reduction in hospital admission.^48^ Furthermore, the SOLIDARITY trial has reported no definite mortality benefit of remdesivir (RR=0·95, CI 0·81-1·11; 301 deaths in 2743 remdesivir group, 303 deaths in the 2708 control group), even following a subgroup analysis for severity of illness.

Preprint results indicating that ivermectin may have a role in COVID-19 prophylaxis have recently been released on ClinicalTrials.gov (NCT04425850, NCT04422561).^53^ These are summarised in Table 2. The IVERCAR study enrolled 229 healthcare workers in Argentina and divided participants into two groups, with the treatment group receiving ivermectin buccal drops and carrageenan nasal spray in addition to personal protective equipment (PPE), and the control group using PPE alone (NCT04425850). After 28 days of follow-up, no participants in the treatment group (n=131) had tested positive to COVID-19 compared to 11 participants in the control group (n=98), (p <0·0001).

These results were mirrored in a randomised controlled trial in Egypt, in which 304 participants who had household contacts with confirmed COVID-19 were randomised to receive ivermectin (200-400 μg/kg on day 1 and day 3) or placebo (NCT04422561). Within the 14-day follow-up period, 7·4% of the treatment group had developed symptoms consistent with COVID-19, compared to 58·4% of the placebo group. PCR results had not been released at the time of writing this review, however even a reduction in symptomatic COVID-19 carries important individual health implications. These studies are in contrast to less encouraging or conflicting results from HCQ prophylaxis studies.^56-58^

Consistent with other prophylaxis reports, a recently released preprint matched case control study on medRxiv that analysed various medications used experimentally as COVID-19 prophylaxis, reported a 73% reduction of COVID-19 in healthcare workers following two doses of ivermectin (OR, 0·27; 95% CI 0·15-0·51).^59^ The ivermectin prophylaxis regimen used in this study consisted of two doses of 300 μg/kg of ivermectin separated by 72 hours.

## FUTURE DIRECTIONS

Evidence is mounting which suggests that ivermectin may be an important drug in the fight against COVID-19. In the midst of a global pandemic the importance of preprint studies has come to the fore; however, we still await the outcome of rigorous peer review. Attributing therapeutic benefit to ivermectin use alone will be challenging, as combination therapies were commonly deployed. Interestingly, the apparent benefits of ivermectin or ivermectin combination therapy against COVID-19 appear to potentially be relevant to all stages of illness, from prophylaxis to treatment of critically ill patients. This may be explained by the multi-pronged effects of ivermectin, which range from direct viral inhibition to immunomodulation to mitigation of cell access, as demonstrated by in vitro, in silico and animal studies. Further immunological work needs to be undertaken to determine the specific mechanism of action of Ivermectin with respect to its anti-inflammatory effects in COVID-19. Ivermectin is widely available, inexpensive, easy to administer and has a wide safety margin. The potential benefits of ivermectin may be enhanced by non-parenteral drug delivery. Three studies have recently published safety data in animal models of inhaled and intranasal ivermectin, reporting that these routes are safe in their respective models.^60-62^ Further research in dosing, routes of administration, synergistic therapies and drug interactions will help inform the safest and most efficacious approach.

## Data Availability

Data referred to in the manuscript is accessible via references included in the "References" section. They include peer-reviewed and pre-print sources

## ACKNOWLEDGEMENTS

Thank you to Dr. Salvatore Fiorenza for valuable discussions and assistance with manuscript proof reading.

